# The Impact of Health Information Technology on Patient Safety

**DOI:** 10.1101/2024.11.11.24317119

**Authors:** Gideon Danso, Obed Owumbornyi Lasim

**Affiliations:** Health Information Officer/ Research Assistant University of Cape Coast,Department of Health Information Management PMB, Cape Coast, Ghana; Academic Lecturer University of Cape Coast, Department of Health Information Management PMB, Cape Coast, Ghana

**Keywords:** Impact, Health Information, Health Information Technology, Patient Safety

## Abstract

**Introduction:** Since the original Institute of Medicine (IOM) report was published there has been an accelerated development and adoption of health information technology with varying degrees of evidence about the impact of health information technology on patient safety. We conclude that health information technology improves patient’s safety by reducing medication errors, reducing adverse drug reactions, and improving compliance to practice guidelines. There should be no doubt that health information technology is an important tool for improving healthcare quality and safety.

**Method:** A cross-sectional survey was conducted among some health care professionals at St. Gregory Catholic Hospital, Gomoa Buduburam. A self-administered questionnaire was used to elicit information on informed consent, confidentiality and medico legal issues. Data collected was analyzed using SPSS version 20.

**Results:** A total of 70 health care workers were enrolled on the study representing 70% response rate. The study revealed about 98% knows that Health Information Technology (HIT) is important in delivering care to patients; and 86% of the respondents indicated that HIT has reduced medical errors. About 82% of the responded that HIT has made health care delivery easier and faster and hence reducing awaiting time. About 90% of the responded that HIT has helped in transferring medical records of patient from one facility to the other in case of emergency.

**Conclusion:** Impact of Health Information Technology on patient safety and their perception are positive. However, regular training on the update and use of Health Information Technology will be necessary in order to ensure continuous improvement of the quality of health care delivery.

## 1.0. CHAPTER ONE

### 1.1 Introduction

Customer expectations never stay the same in a world that is always changing and evolving. A recent example is the pandemic. It had a negative impact on many industries, but healthcare was the worst hit. Providers, healthcare workers, and patients all had to prepare for and accept new ways to deal with a scenario in which physical touch was deemed lethal. Telehealth saved providers and allowed people to consult doctors from the convenience of their own homes. Hospitals have been nudged by technology and connected care to automate workflows and extend healthcare services beyond the hospital’s four walls. It has enhanced processes that previously took a long time and appeared to be unattainable. In the twenty-first century, technological advancements have resulted in considerable changes in healthcare. While the use of electronic health records (EHR) has had a significant influence, other technologies have arisen that are making healthcare more efficient and increasing patient safety and health outcomes. Telehealth and telemedicine, as well as wearable medical gadgets and interoperable systems, are among these advancements. Putting this new technology to work necessitates a strong partnership between health physicians and health Information Technology (IT) professionals, who ensure that the technology is used to support medical personnel and patient safety to the greatest extent possible. Today’s facilities, providers, hospitals, and health companies want powerful and flexible healthcare IT systems that not only fulfill current needs but also prepare them for future prospects.

Because of the application of information technology, health care organizations have claimed a considerable improvement in the quality of care. Information technology allows for easier access to explicit care guidelines, which prevents processing errors and improves care quality. Health Information Technology (HIT) plays a significant role in Care settings and it impacts patient care and patient safety. A Congressional Budget office study stated that EHRs can reduce prescription errors by 95%. Furthermore, last decade saw the adoption of EHR in a major way, across the world. This has led to the major shift, the migration from paper records to EHR has benefitted providers, payers, and patients in many ways. There is no need for searching the paper records, copy and faxing records to other hospitals and providers; make phone calls to discuss the patient’s condition during an emergency, or before inter and intrastate transfers. Information technology makes specific care instructions more accessible, which reduces processing errors and enhances care quality. HIT has a substantial impact on patient care and patient safety in healthcare settings. According to a study conducted by the Congressional Budget Office, EHRs can reduce prescription errors by 95%. Furthermore, the recent decade saw widespread use of electronic health records (EHR) around the world. This has resulted in a huge shift, with the migration from paper records to electronic health records (EHR) benefiting clinicians, payers, and patients in a variety of ways. There’s no need to look through paper records, copy and fax records to other hospitals and doctors, or make phone calls to discuss the patient’s health during an emergency or before interstate or intrastate transfers. It is simple for a physician to send patient data to other providers using modern EHR. Medication errors, adverse drug responses, and noncompliance with practice recommendations are all reduced by health information technology, which enhances patient safety. Without a doubt, technology plays an important role in increasing the quality and safety of healthcare.

HIT is defined as “the application of information processing involving both computer hardware and software that deals with the storage, retrieval, sharing, and use of health care information, data, and knowledge for communication and decision making.”

Health information technology has been rapidly growing and accepting/adopting, with a range of evidence of its impact on patient safety. The aim of this study is to shed light on recent science regarding the impact on patient safety of different health information technology. Health information technology contributes to improve patient safety by minimizing medication errors, reducing the potential for adverse drug reactions, and improving compliance with practice standards/protocols. Health information technology is undoubtedly an ease. As research indicates that there is little evidence of progress/improving patient safety end results in certain innovations, healthcare institutions need to know which information technology they can finance. Patient safety in health care is characterized as the prevention, prevention and enhancement of adverse healthcare effects or injuries. (Patient Safety Dictionary, 2017 [Internet], National Patient Safety Foundation).

In 1999, the Institute of Medicine (IOM) study To Mistake is Human (Kohn LT, et al, 2000) called for the development and evaluation of new technology to reduce medical error, and the subsequent 2001 report crossing the Quality Chasm (Crossing the Quality Chasm, 2001), [Internet]) called for the use of knowledge to increase patient safety. The subsequent 2001 study, crossing the quality chasm, recommended that information technology be used as a critical first step in improving and evolving the healthcare system in order to provide better and safer treatment. The use of data processing, involving the storage, collection, sharing and use of data, data and knowledge for communication and deciding within the area of health care has been described as information technology (Brailer D, 2004) The decade of health information technology, Framework for Strategic Action ([Internet]). Health information technology covers a wide range of technologies, from basic charts to more sophisticated support for decision-making and integration with medical technology. Reducing human mistakes, enhanced clinical quality, promoting patient planning, improving efficiency of practice and monitoring of information over time are just a few ways to improve and improve healthcare thus health information technology. Since the initial IOM study, health information technology has expanded rapidly and been implemented with different levels of evidence on the effects of health information technology on patient safety. The purpose of this study is to explain the latest science on the effect of diverse information systems in health on the safety of patients. This study may be useful for clinicians and health officials when making decisions on the basis of evidence. The impact of health information technology on patient safety and care is as varied as the systems and devices now in use. However, much of it starts with the adoption of EHR by medical facilities in the last decade. The goal of this study is to discuss the most recent research on the impact of various health information systems on patient safety. Clinicians and health policymakers may find this study beneficial when making evidence-based decisions. As diverse as the systems and technologies already in use, the influence of health information technology on patient safety and care is diverse. However, much of it may be traced back to medical facilities’ use of electronic health records (EHR) in the recent decade. These records serve as a central repository for a patient’s medical history, allowing clinical information such as physician notes, test results, and prescription drug information to be shared. EHRs take human elements into account in healthcare, enhancing communication and offering a “single source of truth” on patient health and treatment for improved patient care coordination. Information technology’s effect has widened beyond information visibility and sharing to include the use of data for data-driven patient care. Advanced data analytics is becoming increasingly important in enhancing patient safety.

The coronavirus pandemic has highlighted the value of health data, with analytics algorithms able to construct COVID-19 patient risk profiles in the early days of the pandemic, better identifying those most at risk of lethal effects. With so much technology accessible, healthcare leaders and health IT experts must take a systematic approach to implementing the technology that best suits their needs. According to the Institute for Healthcare Improvement, this works best when three important elements of technology in healthcare are understood.

1. Understanding Needs: Administrators, clinical leaders and IT leaders must come together to identify areas where technology will benefit their specific operation, then set goals for what they want to achieve with that technology.
2. Asking the Right Questions: Both health IT professionals and clinicians must consider every potential tech solution through the lens of how it will impact the work of clinicians, lead to improved patient care and safety, and ultimately, positive health outcomes.
3. Bringing Health IT and Health Clinicians Together: Asking the right questions is dependent on health clinicians and health IT coming together for greater coordination between the work each group does. That’s the best way to get on the same page about how to approach integrating technology. For example, having IT personnel attend clinical meetings could provide greater insight into medical operations.

In many areas, including healthcare, artificial intelligence (AI) and machine learning have vast potential and untapped uses. AI can enhance and expand CDS, according to the same AHRQ report. According to the paper, AI systems now under development can detect complex illnesses like sepsis or other warning signals of patient deterioration. AI and machine learning can also be used in the healthcare industry. Many mundane billing and coding operations may be handled by advanced computers, allowing humans to focus on more complicated tasks. Predictive health analytics is another topic that has emerged in recent years. Advanced systems will be able to suggest the optimum course of therapy for a patient and compare prospective outcomes based on the various options available to clinicians in the future. Technological breakthroughs will shape the future of healthcare. For professionals who want to be on the cutting edge of using technology to improve patient safety and health outcomes, graduate certificate and degree programs in health informatics and healthcare analytics can give the expertise needed to lead in the sector. The majority of obstetrician–gynecologists now use electronic health records. They have quickly acquired traction as a result of the recognition of their potential benefits and government programs that encourage their use. Health information technology’s advantages include the ability to store and retrieve data, as well as the ability to quickly communicate patient information in a legible format, improved medication safety through increased legibility, which may reduce the risk of medication errors, and the ease with which patient information can be retrieved (IT). Technology, among other things, speeds up the flow of information between doctors, lowers the chance of medical errors, enhances drug safety, expands access to medical knowledge, and encourages patient-centered treatment, among other things. Healthcare information technology has the potential to improve and alter health care in a variety of ways, including minimizing human errors, improving health outcomes, and increasing practice efficiency. By automating and implementing medication reminders, using technology to introduce medication alerts and software to manage clinical practice, and improving diagnostic and consultation reports, technology can help to improve patient safety while also improving clinical decision-making, preventing mistakes, and reducing variation in practice. Technology can promote patient-centered care by enhancing contact between clinicians and patients through the use of online portals, SMS, and emails.

Furthermore, it facilitates internet access to medical records, allowing patients to better track their health. Health care practitioners can give better care to their patients if they have a complete and accurate medical record. Electronic health records (EHRs) can help patients’ outcomes by diagnosing diseases, reducing medical errors, and even preventing them.

The potential to improve patient safety exists through the use of medication alerts, clinical flags and reminders, better tracking and reporting of consultations and diagnostic testing, clinical decision support, and the availability of complete patient data. Data gathered through the use of health IT can be used to evaluate the efficacy of therapeutic interventions and have been demonstrated to lead to improvements in the practice of medicine. Alerts can optimize adherence to guidelines and evidence-based care (Classen et al., 2010) Meaningful use of computerized prescriber order entry in Patient Safety [PubMed]). Record uniformity can be designed to reduce practice variations, conduct systematic audits for quality assurance, and optimize evidenced-based care for common conditions (Brokel & Harrison, 2009) Redesigning care processes using an electronic health record: a system’s experience. Health IT is increasing patient engagement as consumers of health care. It allows patients access to their medical records, which helps them to feel more knowledgeable about their conditions and encourages them to actively participate in shared decision making. (The Joint Commission., 2008) Safely implementing health information and converging technologies. Sentinel Event Alert Issue (No. 42. Oakbrook Terrace (IL)). Outside the patient encounter, it can improve follow-up for missed appointments, consultations, and diagnostic testing. A health care provider can search for specific cohorts of patients within a practice to monitor and improve adherence to indicated health care such as mammograms, Pap tests, or measurement of hemoglobin A1c levels. In addition to being able to store and retrieve data, IT can be used to rapidly convey patient information in a legible format, improve medication safety by increasing legibility and thereby potentially reducing medication errors as well as facilitating retrieval of data.

### 1.2. Problem Statement

Patient safety is the most important requirement when providing care to a patient. The primary goal of physicians is to ensure that their patients are treated, safe, and recover from their illnesses while under their care. Every physician’s goal is to provide safety to their patients who visit them. There have been numerous documented errors in the health care setting that have resulted in the death of people seeking medical attention. Medical errors on the part of physicians have resulted in the deaths of illustrious people. Some errors can be easily identified and corrected by using information technology such as EHR and PDMS, for example. Errors are part of everyday life for humans, so physicians are also prone to making mistakes because they are also humans. Adverse drug reactions and noncompliance with practice guidelines are also major issues that endanger patient safety. Failure to administer the appropriate medication in response to the correct diagnosis may result in death. It is critical for physicians to provide the appropriate medications to patients who have been diagnosed with a disease related to the prescribed medications. Ineligibility to write patient prescriptions may cause the pharmacy to give the patient the incorrect medication. Clinical Decision Support Systems (CDSS) can be used to effectively administer prescribed medication to patients while avoiding errors that could result in death. Ineligibility of patient demographics also leads to the loss of lives. For example, if a patient’s age is supposed to be 26years but instead 6years was written, the physician’s prescription will vary considering the age and can affect the patient, eventually leading to death or delay in treatment. The intervention and emergence of health information technology has greatly aided in improving patient safety and lowering the rate of mortality.

### 1.3 Specific Objectives

1. To determine the impact of health information technology on patient safety
2. To examine the usage of health information technology
3. To explore the difficulties in using the health information technology

### 1.4. Hypothesis

In the healthcare setting, health information technology has aided in improving patient safety. This finding will also assist health care practitioners in understanding the value of health information technology, as well as the numerous benefits and ease of use that it brings to patient safety.

- **1.5** Research Questions

1. What impact has the use of health information technology been on patient safety?
2. How do health providers use health information technology often?
3. Has there been difficulties in the usage of health information technology?
- **1.6** Significance of the study

Patient safety is a subset of healthcare and is defined as the avoidance, prevention, and amelioration of adverse outcomes or injuries stemming from the processes of health care. (Patient safety dictionary, 2017) and also help to reduce medical errors and improve patient safety. In this modern Era of technology, the introduction of Health Information Technology (HIT) will help with the solving of problems and improving of patient safety. In 1999 the Institute of Medicine’s (IOM) report “To err is human” called for developing and testing new technologies to reduce medical error, (Kohn, Corrigan, & Donaldson, 2001). This research will assist to determine the impact of health information technology on patient safety at St. Gregory Catholic Hospital, of which this study will help improve patient safety by the use of health information technology among health care professionals such as physicians, nurses and other health care providers on the need to determine how health information technology has impacted patient safety in St. Gregory Catholic hospital.

### 1.7. Scope of the study

This study is based on the impact of health information technology on patient safety, with regards to this, this project work is focused on St. Gregory Catholic Hospital, Buduburam in the Central Region of Ghana.

## 2.0. CHAPTER TWO REVIEW OF THE LITERATURE

### 2.1. Health Information Technology (HIT)

Health Information Technology (HIT) is defined as the application of information processing involving both computer hardware and software that deals with the storage, retrieval, sharing, and use of health care information, data, and knowledge for communication and decision making. The Internet of Healthcare Things (IoHT) and HIT play a vital role in preventing errors, saving time, safety, and alerts. HIT encompasses various technologies ranging from simple charting, advance decision support, and integration with medical technology. HIT presents enormous opportunities for transforming healthcare, some of the examples include:

- Minimizing human errors: we human beings cannot do away with errors as it is part of us. HIT has been designed to give a quick prompt or notification readings whenever it is used and this has made it very easy for humans to track where there is a problem. Hence has minimized human errors.
- Improving clinical outcomes: clinical outcomes are measurable changes in health, function or quality of life that result from our care. Due to HIT, there has been improvement in clinical results and aids in the prevention and disease management.
- Facilitating care coordination: HIT has helped in the facilitation of care coordination among healthcare providers. It has made it very easy for providers to communicate with one another when using HIT in the care of patients.
- Improving practice efficiencies: HIT has helped in the improving of practice efficiencies among health care practitioners.
- Tracking & analyzing data: Due to the storage ability nature of HIT, it has made it easier for healthcare practitioners to track the progress of care of patients and also help in the analyzing of data.

There are several technologies that help in the improvement of patient safety and some of these include the following: Electronic Health Record (EHR), Electronic medical record (EMR), Electronic Physician Order Entry (CPOE), Clinical Decision Support (CDS), E-prescribing, Electronic Sign-out and Hand-off, Universal Product Code Medication Administration (BCMA), and so on.

### 2.2. Electronic Health Records (EHR)

Health Information Management System Society (HIMSS) defines Electronic Health Record; EHR could also be a longitudinal electronic record of patient health information generated by one or more encounters in any health care delivery setting. Included during this information are patient demographics, progress notes, problems, medications, vital signs, past medical record, immunizations, laboratory data, radiology reports etc. Computer based Patient Record Institute (CPRI) now a part of the Healthcare Information and Management Systems Society (HIMSS): identified three key criteria for an EHR; The electronic health record must:

- Integrate data from multiple sources/departments
- Capture data at the purpose of care
- Support caregiver deciding

Health Information Management Systems Society’s;(HIMSSJ) emphasis that:

The EHR automates and streamlines the clinician’s workflow. The EHR has the power to get an entire record of a clinical patient encounter, also as supporting other care related activities directly or indirectly via interface including evidence-based decision support, quality management, and outcomes for reporting. it’s important to notice that an EHR is generated and maintained within an establishment, like a hospital, integrated delivery network, clinic, or physician office.

### 2.3. Electronic Physician Orders and E-Prescribing

Electronic physician orders and e-prescribing are two samples of electronic physician orders and e-prescribing.

The use of electronic or network support to enter physician orders, including prescription orders, employing a computer or mobile device platform is understood as computerized physician order entry. (Computerized Provider Order Entry [Internet] Agency for Healthcare Quality & Research, 2017). Originally designed to reinforce the security of prescription orders, computerized physician order entry systems now leave the electronic ordering of tests, treatments, and consultations. Clinical decision support systems (CDS) are typically incorporated with computerized physician order entry systems to function a mistake prevention tool by advising the prescriber on the specified drug doses, path, and pace of administration. (Nuckols et al., 2013). Furthermore, some CPOE systems may have the potential of alerting the prescriber to any patient allergies, drug-drug or drug-lab interactions, or, in additional advanced systems, interventions that ought to be recommended supported clinical guideline recommendations (example venous thromboembolism prophylaxis). The introduction of a deal with clinical decision support resulted during a substantial reduction in medication errors, consistent with a meta-analysis assessing the efficacy of CPOE to attenuate medication errors and adverse drug events in hospitals. Similarly, reviews of community-based outpatient facilities revealed similar reductions in prescription errors. The utilization of hard-stops in CPOE systems as a measure of forcing feature and error prevention has been investigated and located to achieve success in reducing prescribing errors. The utilization of hard-stops, on the opposite hand, resulted in clinically significant treatment delays the utilization of a stand-alone CPOE within the absence of CDS doesn’t appear to scale back medication errors.

### 2.4. Clinical Decision Support

Clinical decision support provides information and patient-specific information to health care professionals. This information is meant to assist the healthcare provider make a far better decision, and other options are presented to the healthcare professional at appropriate times. Consistent with a Cochrane systematic review, using on-screen reminders for physicians resulted in minor to modest improvements in process adherence, medication ordering, vaccination, laboratory ordering, and clinical results.

A meta-analysis investigated why some CDS systems succeed and improve patient outcomes while others fail, and concluded that CDS systems that provided simple advice were less likely to succeed, whereas CDS systems that required the healthcare provider to justify the rationale for overriding CDS advice were more likely to succeed. The probabilities of success were also higher for CDS systems that provided advice to patients and practitioners at an equivalent time. Furthermore, CDS systems evaluated by their developer instead of third-party developers were more likely to succeed. (Roshanov et al, 2012). Although published research shows that CDS systems improve quality of care and patient safety, the results may vary counting on system design and implementation methods.

### 2.5. Electronic Sign-out and Hand-off Tools

Sign-out or “hand-over” communication refers to the tactic of passing patient-specific information from one caregiver to subsequent, from one team of caregivers to subsequent, or from caregivers to the patient and family so on confirm patient care continuity and safety. (Joint Commission (2014, p.23): The Joint Commission International Accreditation Standards for Hospitals. [Google Scholar]). One of the leading root causes of sentinel events within the US has been identified as a breakdown in patient information handover. (Popovich, 2011) 30-Second Head-to-Toe Tool in Pediatric Nursing: Cultivating Safety in Handoff Communication). Electronic sign-out applications are tools which can be used independently or in conjunction with an electronic medical record to form sure a structured transfer of patient information during provider handoffs. Two systematic reviews (Davis et al., 2013) evaluating the outcomes of electronic tools supporting physician shift-to-shift handoffs concluded that the bulk studies supported using an electronic tool with an improvement within the handover process.

### 2.6. Bar Code Medication Administration

Electronic systems that integrate electronic medication administration records with Universal Product Code technology are mentioned as Universal Product Code medication administration systems. These systems are designed to prevent medication errors by ensuring that the proper medication is given to the proper patient at the proper time. Furthermore, existing barcode systems differ in their level of sophistication. Observational or quasi-experimental studies provide the absolute best level of clinical evidence for this technology. a scientific review of quasi-experimental studies (Leung et al., 2012) discovered that integrating Universal Product Code medication administration with electronic medication administration records can reduce medication administration errors by 50% to 80%. The systematic review, however, didn’t enter detail about whether the included studies were evaluated for the quality of their methodology. The review also stated that there is a scarcity of data on the use of barcode technology in pediatric and outpatient settings, as most studies are conducted in an inpatient adult setting. Another systematic review conducted a meta-analysis of studies involving BCMA and discovered that implementing BCMA resulted during a 57 percent reduction in medication errors. This result, however, should be interpreted with caution because the studies included within the meta-analysis were highly heterogeneous. Although BCMA automates and improves medication administration documentation, there’s only moderate to weak clinical evidence that it’s effective in reducing medication costs. to achieve a conclusion, more thorough research is required. Healthcare organizations must also consider the impact of BCMA implementation on their workflows.

### 2.7. Smart Pumps

Intravenous infusion pumps with drug error prevention tools are mentioned as smart pumps. When the infusion setting is about outside of pre-configured safety limits, this program warns the operator. the only reported randomized controlled trial on the effect of smart pumps on medication safety found no statistical difference between turning on the selection support function on or off the smart pump. The authors clarified that this was due partially to healthcare providers’ inadequate compliance with infusion procedures. Smart pumps can minimize programing errors, but they’re doing not eradicate them, according to a scientific review of quasi-experimental studies. Hard limits were also found to be simpler than soft limits in preventing drug errors, according to the report. The high rate of sentimental limit override explained this. To draw a conclusion about the effectiveness of smart pumps in reducing medication errors and improving patient safety, further research is required.

### 2.8. Automated Medication Dispensing Technology

ADCs (automated dispensing cabinets) are electronic drug cabinets that store medication at the aim of care and permit for managed dispensing and delivery monitoring. Within the 1980s, hospitals began using automated dispensing cabinets, which have since expanded to incorporate more advanced software and interactive interfaces to synthesize high-risk measures within the drug dispensing method. Automated drug dispensing cabinets are successfully used as a drugs control tool to help simplify the medication dispensing process by reducing central pharmacy workload and keeping better track of medication dispensing and patient billing. The effect of ADC on patient safety is minimal, as there’s only one reported controlled trial, which showed that using ADC decreased drug errors by 28 percent (p0.05) during a hospital critical care unit (RR: 0.7; NNT: 4). the majority of the errors that were minimized were planning errors, according to a thorough review. The automated dispensing system had no effect on the quantity of errors that caused damage. In critical care settings, automated dispensing cabinets appear to attenuate drug preparation errors. Despite the high standard of data, it’s only applicable to critical care environments. To draw a conclusion about the effect of ADC on drug protection in to, further controlled studies are required.

### 2.9. Patient electronic portals

A patient portal could also be a secure online application that allows patients to access their personal health information and communicate with their care provider in two ways employing a computer or a mobile device. Numerous studies have shown that patient portals improve preventive care, disease awareness, and self-management outcomes. There is no evidence, however, that they improve patient safety outcomes.

### 2.10. Telemedicine

Telemedicine is defined because the utilization of telecommunication technologies to facilitate communication between patients and providers. Communication is often synchronous, with real-time 2-way video communication, or asynchronous, with patient clinical information transmitted asynchronously. Telemedicine also can provide health information collected remotely from medical devices or personal mobile devices, additionally to communication. This information could even be used to monitor, track, or change the behavior of patients.

#### 2.10.1 Synchronous Telemedicine

A virtual visit could also be a two-way audio/video communication between a healthcare provider and a patient that takes place in real time. Numerous systematic reviews are conducted to research the impact of virtual visits on patient outcomes in critical care, chronic disease care, and psychiatric care. In terms of specific clinical outcomes, all have shown that telemedicine is as effective as face-to-face care, but there’s limited evidence regarding patient safety outcomes. An e-consultation could also be a secure communication platform-based transmission between the patient’s medical aid clinician and a specialist. The technology provides the specialist with guidance on patient management without reference to the patient. Efficacy and safety in e-consultations are limited, but studies have shown that e-consultations can reduce the wait times for specialist appointments and opinions for patients.

### 2.11. Remote Patient Monitoring

Community-based studies the remote monitoring of patients (telemonitoring) demonstrate that it enhances patient outcomes for chronic conditions, e.g. heart failure, stroke, COPD, asthma, and high pressure sign. Patient data administration system (PDMS) could also be a system where the data from bedside medical equipment is automatically collected (namely patient monitor, ventilator, intravenous pump, then forth). The data is submitted to help healthcare providers interpret the data and subsequently restructured. PDMS has been integrated with clinical decision support and electronic medical records in recently progressed integration. A systemic review examined the clinical implications of PDMS and determined that these systems spent longer on direct patient care by reducing the time spent on charts. Furthermore, PDMS systems reduced error occurrence (medication errors, ventilator incidents, intravenous incidents, and other incidents). The review found that the clinical outcomes improved by integrating PDMS with a clinical decision network were reported in 2 articles. Research shows that telemedicine technology appears to reinforce clinical outcomes surely medical conditions and appears to make health services more accessible, encouraging collaboration between patients and physicians. Besides the limited evidence on PDMS, there seems to be little clarity about the impact telemedicine on patient safety.

### 2.12. Electronic Incident Reporting

Electronic incident reporting systems are web-based systems that enable healthcare providers involved in security events to report incidents on a voluntary basis. The Electronic Health Record (EHR) allows such systems to take in data and automatically detect negative events using trigger tools. the next potential benefits are given to electronic incident reporting systems: the standardization of reporting structures, a consistent workflow for action by incident, quick detection and triggering events, while automating data entry and analysis. Published research shows that the reporting frequency has increased considerably among healthcare organizations which have moved into the electronic reporting system. Clinical processes may improve incident reporting systems, but there’s little proof that electronic reporting systems eventually reduce medical errors.

## 3.0. CHAPTER THREE METHODS OF THE STUDY

### 3.1. Introduction

This chapter explores the research procedure and methods that are used in the study. It identifies the research approach and design and justifies their usage, the population, sampling procedures, research instruments, data gathering process and administration of the study.

### 3.2. Research Approach and Design

A research approach is viewed as a method or strategy for collecting and analyzing data required to answer a research question. In research studies, a researcher can adopt any of the three (3) research approaches, thus: quantitative, qualitative or mixed (Cohen et al, 2011). The study follows a cross-sectional design using the mixed method approach to quantify the knowledge of Healthcare Professionals on the impact of Health Information Technology on patient safety. Cross sectional survey was used since it helps to collect data on the whole sample population at one specific point in time.

### 3.3. Study Area

This study was conducted at Saint Gregory Catholic Hospital, Kasoa Budumburam. The hospital started as a clinic in 1990 to receive Liberia Refugees from the civil war. Apam catholic hospital under the directives of his eminent cardinal Peter Appiah Turkson started a medical rehabilitation for the refugees in 2002, the archdiocese of Cape Coast asked Apam Catholic hospital to reopen the clinic to help with health delivery in the camp. With regards to Catchment Area/population, the facility caters for clients far and beyond. Their immediate and often beneficiaries’ stretches Nyanyano through Kasoa to Bawjawse and from Kasoa to Okyereko. Beyond these, our clients come from as far as Pokuase. Officially, the hospital operates by the certified Population of Budumburam 50,855(i.e., 2010 population census). St. Gregory Catholic currently operate on the following units; Emergency, Anti-natal care, Pharmacy, Laboratory, Radiology, ENT, Wards for Male, Female and Children, Theatre, Out Patient Department, Quality Improvement, Health Information etc.

### 3.4. Target Population

A population is defined as a group of all objects that exhibit one or more desired characteristics (Rahawarin & Arikunto, 2015). A population according to Brantlinger et al. (2005), is a group to which a researcher would like the findings of the study to be generalizable. The target population for this study was healthcare professionals who use Information Technology (IT) in providing care to patients at St. Gregory Catholic Hospital.

### 3.5. Sample and Sampling Procedure

The study was conducted on only healthcare professionals using IT to provide care to patients at St. Gregory Catholic Hospital for accurate results on impact of HIT on patient safety. The sample size for this study was made up of 70 respondents which were selected through simple random sampling procedure. The population of healthcare professionals at St. Gregory Catholic Hospital was almost 100 as at when the study was conducted. The simple random sampling procedure was used to select 70 healthcare professionals representing about 70% of the population.

#### 3.5.1. Sample size determination

By using StatCalc function in Epi-Info software version 1.4.3 and at a confidence level of 90% and a margin of error 10%, a sample size of 70 was selected from a population of 100 at the time at which data collection was taking place at St. Gregory Catholic Hospital.

### 3.6. Data Collection Method and Data Analysis

A structured researcher administered questionnaire was used to collect data from respondents in the form of an interview. Copies of the questionnaire were given out to the health care professionals at the various departments. I happened to do my internship at the health information department so it made it less stressful to get to access to the clinicians. All respondents were anticipated to give their best cooperation, as the information on the questionnaire is all about the things that revolve around the study. With this, enough time was taken to clarify on the things stated in the research questionnaire.

The questionnaire comprised of three sections excluding the data collected from respondents about their demography:

i. SECTION A

In this section, information on the rating of impact of HIT were obtained. Respondents were asked to choose from the five Likert items below: **strongly agree, agree, neutral, disagree, strongly disagree.**

ii. SECTION B

Information concerning the level of usage of Health Information Technology were obtained in this section. The respondents were asked questions/statements to tick with same options as in section A.

iii. SECTION C

This section collects information explores the challenges in the usage of HIT. The respondents were asked questions/statements to tick with same options as in section A.

### 3.7. Data Analysis and Presentation

Analyses for this study was performed using the Statistical Package for Social Sciences (SPSS) version 26.0 and was organized and edited in Excel spreadsheet. Also, descriptive statistics was used to summarize results and presented them in tables and charts.

### 3.8. Ethical Considerations

Permission to carry out the study was obtained from the administration of St. Gregory Catholic Hospital. Verbal content was obtained from respondents in the language they understand. Respondent assured of confidentiality and the right to withdraw from the study at any point in time before questionnaires were administered.

## 4.0. CHAPTER FOUR RESULTS AND DISCUSSION

### 4.1. Introduction

This chapter focuses on the presentation of the results, analysis and discussion of the results of the study. These are discussed and presented according to the main research questions stated to assist in the study in chapter one. Frequencies and percentages were used for the data analyses. The questionnaires developed were to be distributed to only 70 respondents (sampled population) of the total population (100) during the data collection. However, 70 out of 70 respondents targeted, answered the questions. It represents 70% of the sampled population. Therefore, the aim was achieved. The 70 respondents used for the study were only healthcare professionals of St. Gregory Catholic Hospital, Budumburam in the Central Region, Ghana. The main focus of the study was to know the impact of Health Information Technology on patient safety among healthcare professionals. This chapter is grouped into four sections, in relation to the research questions as well as the information sought from respondents. The results of the study are subsequently discussed

**Key**

S.A = Strongly Agree A = Agree U = Uncertain

S.D = Strongly Disagree D = Disagree F = Frequency

**Table A:**
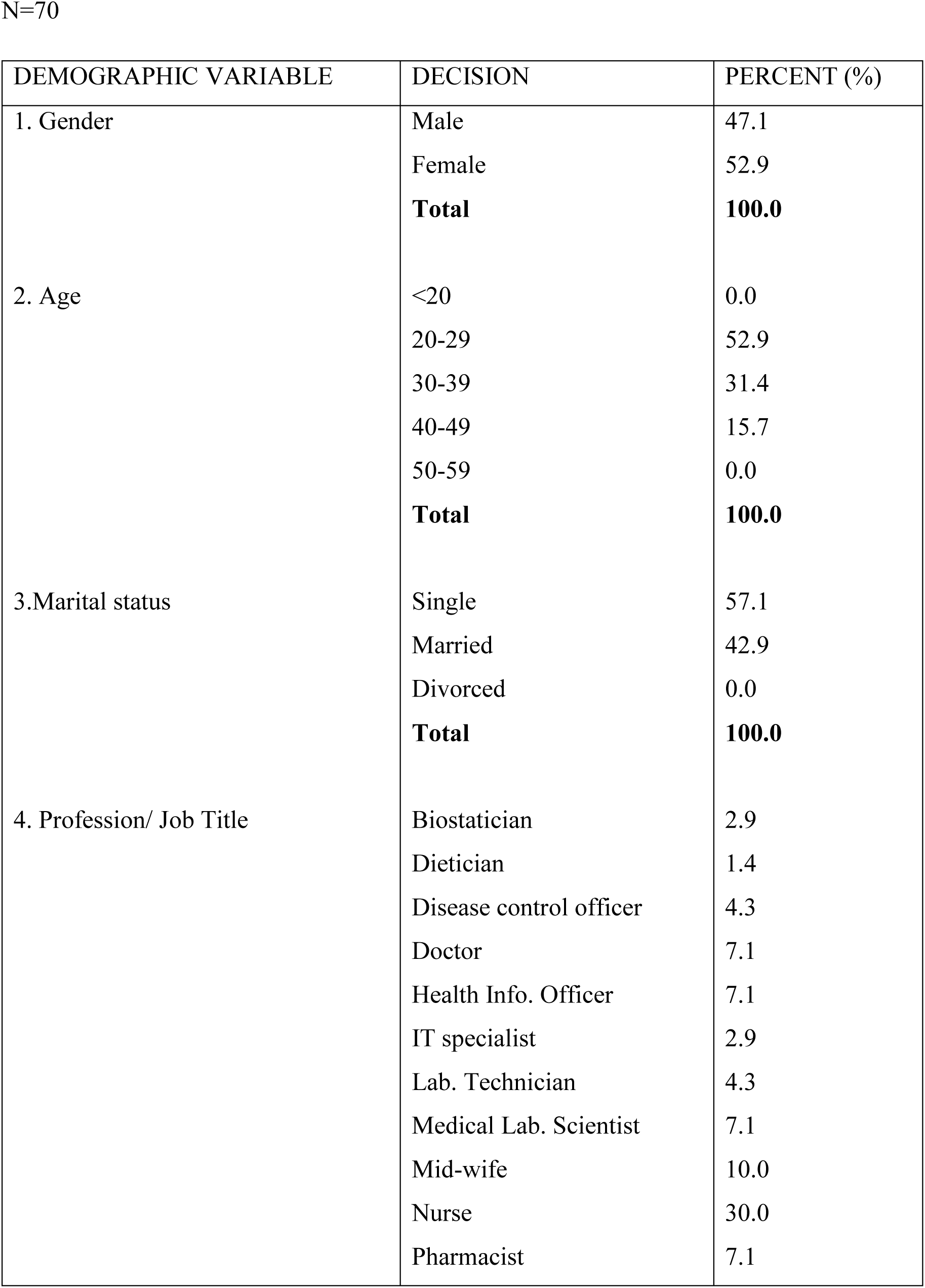
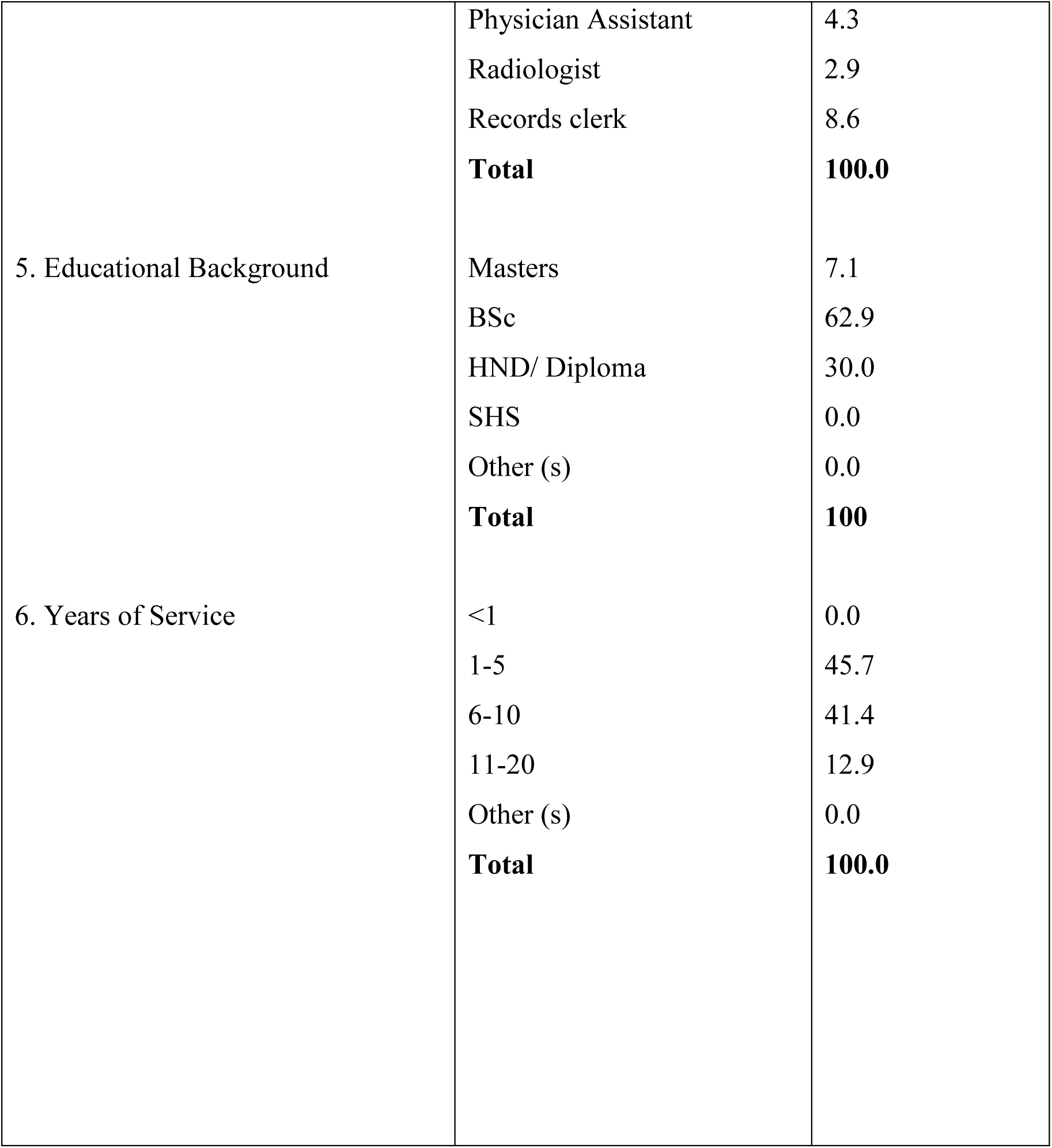
The Demographics, Decision and Percentage of results.

### 4.2. Analysis of table A

The part I of the questionnaire was used to collect data regarding the social-demographic characteristics of the participants. It contained six questions which include gender, age, marital status, profession, educational background and years of service (work experience). Gender was coded as (Male = 1 and Female = 2), age was recorded as (below 20 years = 1, 20-29 years = 2, 30-39 years = 3, 40-49 years = 4, 50-59 years =5 and 60 years and above = 6), marital status was coded as (Single = 1, Married = 2 and Divorced = 3), the profession was also coded, educational background was coded as (Masters = 1, BSc = 2, HND/Diploma = 3, SHS = 4, and other (s) = 5), finally, years of service was recorded as (less than 1 year = 1, 1-5 years = 2, 6-10 years =3, 11-20 years = 4, 16-20 years = 5 and others = 6).

From Table 4.1 above, it was shown that a total of 70 respondents were involved in the research, 33 (47.1%) of the respondents were males and the 37 (52.9%) of the remaining were females. This informs the perception that the healthcare sector is female dominated. Majority of the respondents (52.9%) were between the ages of 20-29 years, (31.4%) of the respondents were between the ages of 30-39 years, (15.7%) of the respondents were between the ages of 40-49 years, none of the respondents were between the ages of 50-59 years, 60 years and above as well as 20 years and below, therefore recording (0.0%) each. This clearly depicts that large number of the healthcare professionals are youthful or young adults. Out of the total number of respondents who took part in the research, 40 of the respondents representing the majority with 57.1% were single, 30 of the respondents with 42.9% were married, there were no case of divorced recorded. 21 (30.0%) representing majority were nurses, 7 (10.0%) of the respondents were midwives, 6 (8.6%) of the respondents were Records clerk, however, some of the respondents who were doctors, pharmacist, medical laboratory technologists/scientists, health info. officer obtained 5 (7.1%) each. Again, disease control officer, lab technicians and physician assistants (PA) recorded 3 (4.3%) each, lastly enough, other respondents including biostatistian, IT specialists, radiologist obtained 2 (2.9%) and dietician obtained 1 (1.4%) respectively. It is worth noting that most the respondents were nurses which imply that there are more nurses in St. Gregory Catholic Hospital, Buduburam than the rest of the healthcare professionals such as the allied health, doctors and other health professionals. It was observed that 62.9% of the respondents had first degrees, 30.0% possessed HND/ Diploma, and 7.1% of them had masters. 32 (45.7%) of the respondents had one to five years (1-5) work of experience, 29 (41.4%) of the respondents had six to ten years (6-10) work of experience, 9 (12.9%) of the respondents had eleven to twenty years (11-20) work of experience. It was observed that very few of the healthcare professionals employed in St. Gregory Catholic Hospital, Buduburam have been working at the facility for the past eleven to twenty years.

Part II to Part IV of the study sought to analyze the responses of the participants based on the three specific objectives stated previously in chapter one. The first objective sought to determine the impact of health information technology on patient safety. The second objective was to examine the usage of health information technology and finally, the third objective sought to explore the difficulties in using the health information technology.

### 4.3. To determine the impact of health information technology on patient safety

Research question one: What impact has health information technology been of good use on patient safety?

The Part II has three tables, A, B and C, which display the responses of the healthcare professionals in St. Gregory Catholic Hospital, Buduburam concerning how to determine the impact of health information technology on patient safety.

Therefore, the tables below show the responses to determine the impact of health information technology on patient safety among healthcare professionals in St. Gregory Catholic Hospital, Buduburam.

**TABLE B:** Determining the impact of health information technology on patient safety.

| STATEMENT | DECISION | PERCENT (%) |
| --- | --- | --- |
| 7.Health Information Technology is important in healthcare delivery | Strongly Disagree | 0.0 |
|  | Disagree | 0.0 |
|  | Uncertain | 0.0 |
|  | Agree | 72.9 |
|  | Strongly Agree | 27.1 |
|  | **Total** | **100.0** |
| 8. Do you believe HIT has made operations in the hospital setting easier and faster | Strongly Disagree | 0.0 |
|  | Disagree | 0.0 |
|  | Uncertain | 0.0 |
|  | Agree | 92.9 |
|  | Strongly Agree | 7.1 |
|  | **Total** | **100.0** |
| 9.Has HIT reduce medical errors? | Strongly Disagree | 0.0 |
|  | Disagree | 0.0 |
|  | Uncertain | 0.0 |
|  | Agree | 88.6 |
|  | Strongly Agree | 11.4 |
|  | **Total** | **100.0** |
| 10.HIT has helped to reduce patient waiting time | Strongly Disagree | 0.0 |
|  | Disagree | 1.4 |
|  | Uncertain | 0.0 |
|  | Agree | 97.1 |
|  | Strongly Agree | 1.4 |
|  | **Total** | **100.0** |
| 11. HIT has made it possible to transfer the records of patients from one facility to another | Strongly Disagree | 0.0 |
|  | Disagree | 0.0 |
|  | Uncertain | 0.0 |
|  | Agree | 94.3 |
|  | Strongly Agree | 5.7 |
|  | **Total** | **100.0** |

### 4.4. Analysis of Table B

From the above ***Table B***, it was noticed that 70 out of the 70(100%) respondents who took part in the study answered **statement 7** with 51(72.9%) of strongly agree response, 19(27.1%) of agree response and none responded for strongly disagree, disagree and uncertain responses respectively. Therefore, majority of the respondent selected strongly agree indicating that HIT is important in healthcare delivery. Also from **statement 8**, 65(92.9%) responded with Agree while 5(7.1%) responded with strongly agree and none responded for strongly disagree, disagree and uncertain respectively. Therefore, majority of the respondent selected agree indicating that they believe HIT has made operation in the hospital setting easier and faster. Moreover, **statement 9** recorded 62(88.6%) with Agree while 8(11.4%) recorded with Strongly agree and none responded for strongly disagree, disagree and uncertain responses respectively. Therefore, majority of the respondent selected agree indicating that indeed HIT has reduced medical error. Meanwhile, **statement 10** indicated that 68(97.1%) of respondents selected Agree and 1(1.4%) responded with Disagree and Strongly agree respectively. Therefore, majority of the respondent selected Agree indicating that most of the respondent agree that HIT has help to reduce patient waiting time. Moreover, **statement 11** indicated that 66(94.3%) of the respondents selected Agree, 4(5.7%) with strongly agree response none for strongly disagree, disagree and uncertain respectively. Therefore, majority of the respondent Agreed that HIT has made it possible to transfer the records of patients from one facility to another.

**TABLE C:** Examining the level of usage of health information technology (HIT) by Healthcare Professionals.

| STATEMENT | DECISION | PERCENT (%) |
| --- | --- | --- |
| 12. HIT has made it possible to identify and search for patient records without difficulty | Strongly Disagree | 0.0 |
|  | Disagree | 0.0 |
|  | Uncertain | 5.7 |
|  | Agree | 85.7 |
|  | Strongly Agree | 8.6 |
|  | **Total** | **100.0** |
| 13. HIT has helped patients to book appointment with their physician at home without necessarily walking to the facility | Strongly Disagree | 1.4 |
|  | Disagree | 0.0 |
|  | Uncertain | 37.1 |
|  | Agree | 60.0 |
|  | Strongly Agree | 1.4 |
|  | **Total** | **100.0** |
| 14. HIT such as Clinical Decision Support Systems (CDSS) has made it easy for physicians to make clinical decisions when delivering care to patients | Strongly Disagree | 0.0 |
|  | Disagree | 0.0 |
|  | Uncertain | 0.0 |
|  | Agree | 98.6 |
|  | Strongly Agree | 1.4 |
|  | **Total** | **100.0** |
| 15. Patients are able to view their medical history from home by the use of patient portals emerged from HIT | Strongly Disagree | 0.0 |
|  | Disagree | 74.3 |
|  | Uncertain | 8.6 |
|  | Agree | 14.3 |
|  | Strongly Agree | 2.9 |
|  | **Total** | **100.0** |
| 16. HIT has made it possible to know the sex of a child before delivery | Strongly Disagree | 0.0 |
|  | Disagree | 1.4 |
|  | Uncertain | 0.0 |
|  | Agree | 71.4 |
|  | Strongly Agree | 27.1 |
|  | **Total** | **100.0** |

### 4.5. Analysis of Table C

From the above ***Table C***, it was noticed that 70 out of the 70(100%) respondents who took part in the study answered **statement 12** with 60(85.7%) of Agree response, 6(8.6%) of Strongly Agree response, 4(5.7%) of Uncertain and none responded for strongly disagree and disagree responses respectively. Therefore, majority of the respondent selected Agree indicating that HIT has made it possible to search and identify patient information without difficulty. Also from **statement 13**, 42(60%) responded with Agree while 26(37.1%) responded with Uncertain and 1(1.4%) for both Strongly Disagree and Strongly Agree respectively. none responded Disagree. Therefore, majority of the respondent selected Agree indicating that HIT has helped patient to book appointment with their physicians without necessarily walking to the facility. Moreover, **statement 14** recorded 69(98.6%) with Agree while 1(4.1%) recorded with Strongly agree and none responded for strongly disagree, disagree and uncertain responses respectively. Therefore, majority of the respondent selected Agree indicating that indeed HIT such as Clinical Decision Support Systems (CDSS) has made it easy for physicians to make clinical decisions when delivering care to patients. Meanwhile, **statement 15** indicated that 52(74.3%) of respondents selected Disagree and 10(14.3%) responded with Agree, 6(8.6) with Uncertain and 2(2.9) with Strongly agree and none for Strongly Disagree respectively. Therefore, majority of the respondent selected Disagree indicating that most of the respondent Disagree that Patients are able to view their medical history from home by the use of patient portals emerged from HIT. Moreover, **statement 16** indicated that 50(71.4%) of the respondents selected Agree, 19(27.1%) with strongly agree response none for strongly disagree,1(1.4%) disagree and uncertain respectively. Therefore, majority of the respondent Agreed that HIT has made it possible to know the sex of an unborn child before delivery.

**Table D:**
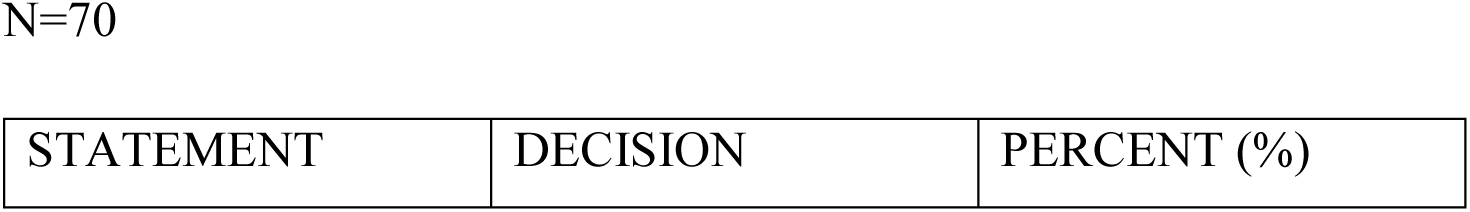
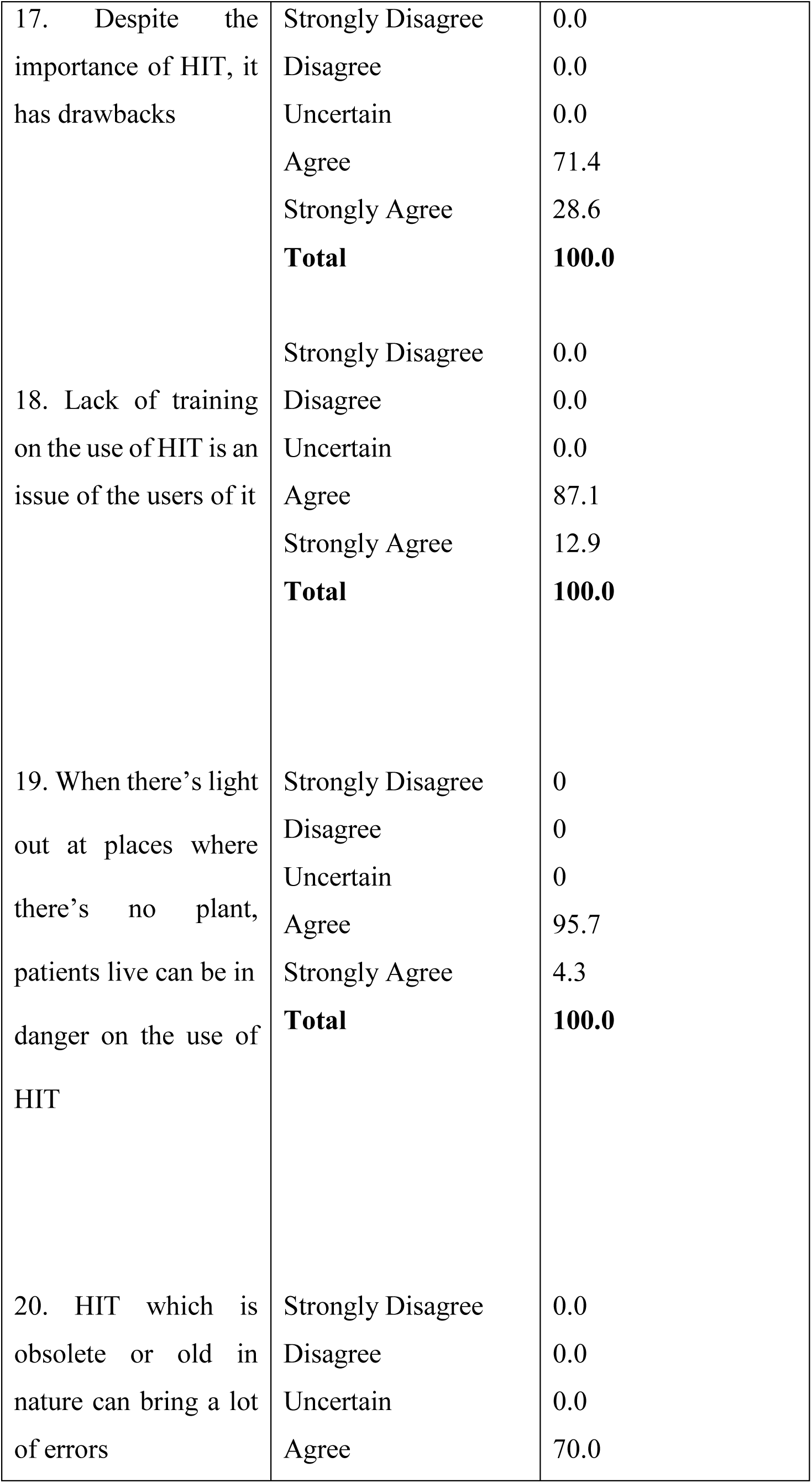
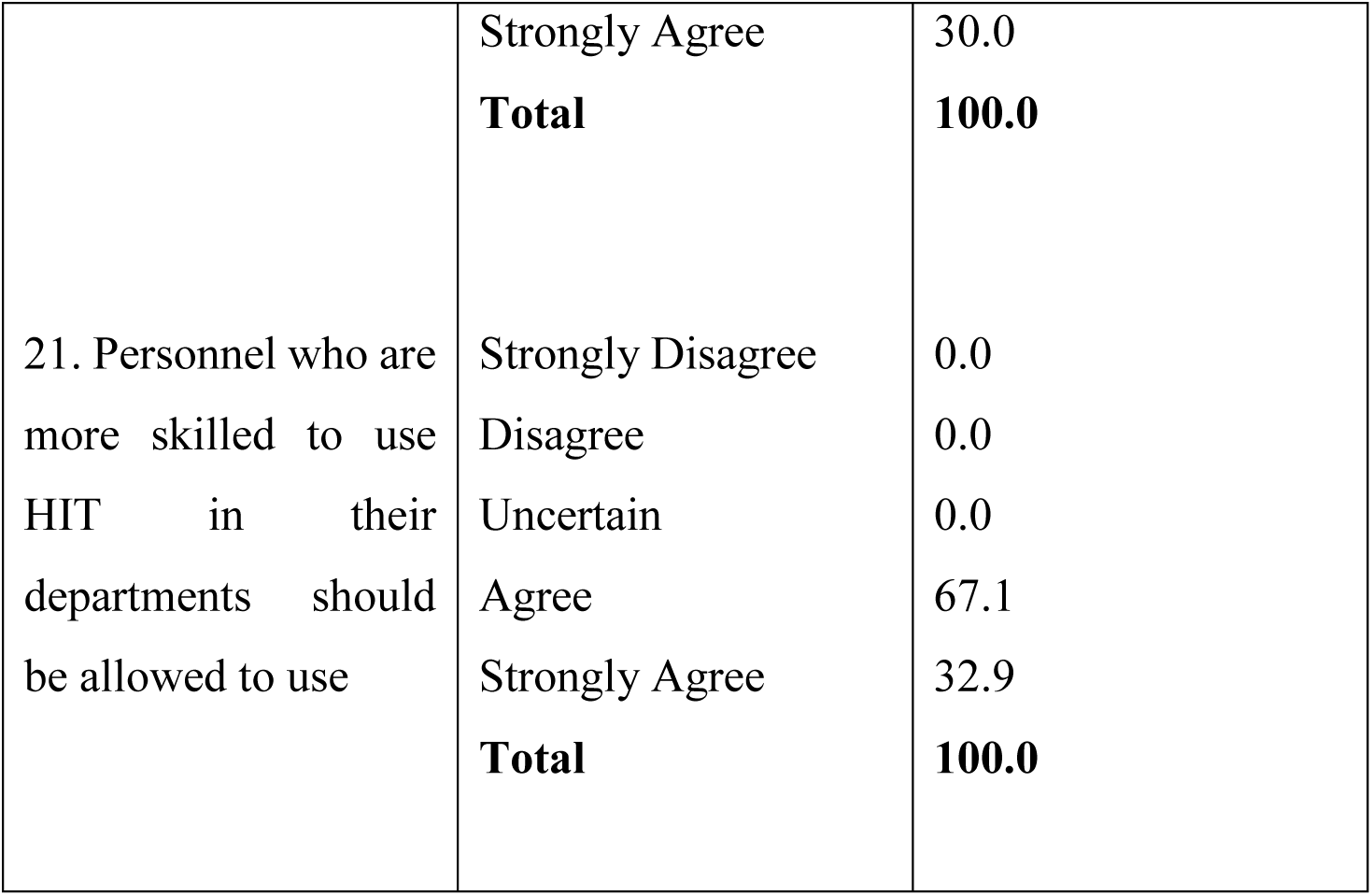
Exploring the Challenges in Using Health Information Technology (Hit) By Health Care Professionals.

### Analysis of Table D

From the above ***Table D***, it was noticed that 70 out of the 70(100%) respondents who took part in the study answered **statement 17** with 50(71.4%) of Agree response, 20(28.6%) of Strongly Agree response, none responded for strongly disagree, uncertain and disagree responses respectively. Therefore, majority of the respondent selected Agree indicating that despite the importance of HIT, it has drawbacks. Also **statement 18** recorded 61(87.1%) with Agree while 9(12.9%) recorded with Strongly agree and none responded for strongly disagree, disagree and uncertain responses respectively. Therefore, majority of the respondent selected Agree indicating that Lack of training on the use of HIT is an issue of the users of it. Moreover, **statement 19** recorded 67(95.7%) with Agree while 3(4.3%) recorded with Strongly agree and none responded for strongly disagree, disagree and uncertain responses respectively. Therefore, majority of the respondent selected Agree indicating that when there’s light out at places where there’s no plant, patients live can be in danger on the use of HIT. More so, **statement 20** recorded 63(70.0%) with Agree while 7(30.0%) recorded with Strongly agree and none responded for strongly disagree, disagree and uncertain responses respectively. Therefore, majority of the respondent selected Agree indicating that HIT which is obsolete or old in nature can bring a lot of errors. Again, **statement 21** recorded 47(67.1%) with Agree while 23(32.9%) recorded with Strongly agree and none responded for strongly disagree, disagree and uncertain responses respectively. Therefore, majority of the respondent selected Agree indicating that Personnel who are more skilled to use HIT in their departments should be allowed to use.

### 4.6 DISCUSSION

A cross-sectional study was carried out at St. Gregory Catholic Hospital, Buduburam to determine the impact of health information technology on patient safety. The study involved data collection in the health facility.

#### 4.6.1 Research Question One: What impact has the use of health information technology been on patient safety?

The impact of health information technology on patient safety and care is as varied as the systems and devices now in use. However, much of it starts with the adoption of EHR by medical facilities in the last decade. These records provide a central repository of a patient’s medical history, allowing for the sharing of clinical information, including physician notes, test results, and information on prescription drugs. EHR’s take into account the impact of human factors in health care improving communication and providing “one source of truth” on patient health and treatment for better coordination of the patient care process. The question sought to investigate what impact on the use of health information technology has been on patient safety. From the result, it was identified most health care professionals agree that the impact of health information technology is positive. The response rate to positive impact on importance of health information technology was about 95% among the respondents but when asked how efficiently used, only 92.3% rate was positive; this could be due to how frequently the health information technology is used in the facility. The above findings agree with a study conducted by Yasser K. Alotaibi et al, (2017), which revealed that There should be no doubt that health information technology is an important tool for improving healthcare quality and safety (Yasser K. Alotaibi et al, 2017). Irrespective of the importance of health information tool, healthcare organizations need to be selective in which technology to invest in to bring more quality of care and as the saying goes “a quality tool bring the quality output”. Hence health information technology has been positive on the impact of patient safety.

#### 4.6.2 Research Question Two: How do health providers use health information technology often?

Health Information Technology (HIT) improves patient safety by reducing medication errors, reducing adverse drug reactions and improving compliance to practice guidelines. The usage of Health Information Technology (HIT) is very important in achieving its benefits and outcomes. The more health information technology is used, the effectiveness and improvement it has on patient safety. From the result, it was identified most health care professionals agree that the frequent use of health information technology is important and help in patient care delivery and patient as well. The response rate to how to search for patient records in case of emergency of health information technology was about 97% among the respondents but when asked how it can be used to book appointment, only 33.3% rate was positive, 60% was uncertain; this could be due to the knowledge and idea of how to use health information technology by patients and I believe educating these patients on how to use the Health Information Technology to book appointment rather than using their time wisely by the care providers whenever they visit the facility for care or for another purpose. The above findings agree with a study conducted by Yasser K. Alotaibi et al, (2017), who revealed that health information technology cannot be a success if both patients and health care practitioners don’t important part. (Yasser K. Alotaibi et al, 2017). With this statement, patients are to also play their part in the success of Health Information Technology. The healthcare practitioners are help with this task by educating the patients on some of the way they can also use the health information technology to improve patient safety. Wearable gadgets and equally other devices to improve patient safety are to be in mind and help to improve care and safety to patients as well. These devices can also help the patients to book appointments with their healthcare practitioners without necessarily going to the facility but rather sit at the comfort of their home and select the date with their provider agreed to visit the facility.

#### 4.6.3 Research Question Three: Has there been difficulties in the usage of health information technology?

In the 2013 list of hazards by ECRI Institute, four of the top ten hazards were directly related to health IT. From the result, it was identified most health care professionals agree that even though health information technology is important in-patient safety, it has drawbacks as well. The response rate to health information technology has drawback is about 85% of the respondents but lack of training on the use of health information technology is an issue among the health care providers is 87% of response rate. The above findings agree with a study conducted by Joint Commission International Accreditation Standards for Hospitals. The Joint Commission. 2014:23 which revealed that “Technology-related adverse events can be associated with all components of a comprehensive technology system and may involve errors of either commission or omission. These unintended adverse events typically stem from human-machine interfaces or organization/system design” (Joint Commission International Accreditation Standards for Hospitals. The Joint Commission. 2014:23.) The use of alerts to warn health care providers of potential problems is a powerful tool. However, alerts top the list of 2013 health IT hazards because the sheer volume of them is causing alert fatigue. This issue is complex and requires individualization within each facility. Developing systems to manage alerts, establish levels of importance, and make them unambiguous is a critical patient safety priority.

## 5.0. CHAPTER FIVE

## SUMMARY, CONCLUSION AND RECOMMENDATIONS

### 5.1. Introduction

This chapter is concerned with the summary, findings, conclusions and recommendations of the study.

### 5.2. Summary

The purpose of the study was motivated by the need to assess the impact of health information technology on patient safety at St. Gregory Catholic Hospital, Buduburam. Three research questions were asked for the study of 70 health care professionals at St. Gregory Catholic Hospital. A 21-statement self-developed questionnaire was used as the main instrument for data collection for the study. Data generated were converted to frequency counts and percentage analysis.

### 5.3. Findings

The main findings of the study are as follows:

- Most people in the world tend to accept technology as part of life and gaining public attention. Health Information Technology which is a type of technology is believed to have solve most problems relating to health matters.
- Most health care professionals are happy with the invention of health information technology and have knowledge about it which makes their work easier and faster.
- The success of a health system is dependent on personnel who have the necessary knowledge, skills and attitudes toward procedures and guidelines to help improve patient safety through the use of health information technology.
- Health information Technology has been designed to solve medical relating issues but the right health care professionals are to be those in charge of these technology to ensure patient safety when delivering care to the patients.
- Not only has health information technology helped solved problems relating to health issues but it has also help to reduce medical errors which can be on the part of the healthcare professionals themselves or less knowledge regarding the subject matter.

### 5.4. Conclusion

The study discovered the importance and relevance of health information technology on patient safety and how the technology can be used effectively by healthcare professionals to successfully deliver care to patients under their care. Health information technology improves patient safety by reducing medication errors, reducing adverse drug reactions and improving compliance to practice guidelines. There should be no doubt that health information technology is an important tool for improving healthcare quality and safety, but healthcare organizations need to be selective in which technology to invest in, as literature shows that some technologies have limited evidence in improving patient safety outcomes. Health IT has become an integral part of the practice of medicine. As with any new technology, health IT brings many potential benefits and as well as potential concerns. The current literature to date, reflects outcomes at single sites or institutions. National estimates are extrapolations from these single-site studies. As the implementation and use of health IT systems increase, it is important to keep patient safety and quality as a major focus. Based on the findings to the study, majority (95%) of the health care professionals are aware that health information technology is important to patient care delivery. Most of them (97%) accept that health information technology has help to reduce medical errors which was a problem some years back. About (85.7%) accepted that health information technology has helped in retrieving patients records faster in case of emergency rather taking a longer time in searching of the patient record whose life is in danger and can lose the life. Time, they say is a precious resource to mankind and therefore (84%) of the healthcare professionals accepts that health information has help to save time and reduce patient waiting time which promotes productivity and enhances patient care and safety. In all, it is believed that health information technology has really positively impact patient safety and therefore has enhance quality care to patient seeking medical attention.

### 5.5. Limitations

The limitation to the study was that there were less trained healthcare professionals at the facility making it difficult to use health information technology in their absence while they are away and also ineffective use by those who are not trained. Thus, those who are not trained to use the health information technology who for some reasons are forced to use it will make errors because they lack the skills in the usage of health information technology.

### 5.6. Recommendations

Though health information technology has really helped in providing care to improve patient safety, there’s the need to take note of some key ways to enable the smooth function and outcome of the health information technology to help deliver care to patients to improve patient safety. The following are some of the ways which can be taken notice to help the smooth functioning of the health information technology:

- Training on the use of common health information technology should be given to all healthcare professionals to enable them to successful deliver care to patients when there’s the need to or when the right people to carry out that specific task are not available. Hence training is a vital element for the use of health information technology. Talking about training, let’s take note of the cost of trainings and also the right people to successfully train the healthcare professionals to fully understand the trainings and to be able to use the technological device very well and help to deliver care and also improve on patient safety. Without trainings healthcare professionals, the objective on the use of health information technology cannot be successfully achieve and, in the end, health information technology won’t help to deliver the needed care to patients.
- Frequent update checks on devices or technology used to deliver care to patients. These technology devices used to deliver care to patients sometimes fail or don’t respond at all and in the end brings about errors when giving care to patients. I suggest these information technologies should be frequently checked or serviced at least once every month to avoid errors which could lead to problem on the part of patient. Also, applications on these technological devices should be updated frequently to match the modern era of technology. Not only the updating of device applications to successfully work on these devices but also the addition of newly improved software programs can be installed on these devices to successfully serve its purpose.

## <u>QUESTIONNAIRE ON IMPACT OF HEALTH INFORMATION</u> <u>TECHNOLOGY ON PATIENT SAFETY AMONG HEALTHCARE</u> <u>PROFESSIONALS AT SAINT GREGORY CATHOLIC HOSPITAL-</u> <u>BUDUBURAM</u>

Dear Respondent,

This questionnaire aims to know the impact of health information technology on patient safety among healthcare professionals in St. Gregory Catholic Hospital, Buduburam.

All healthcare professionals within the facility are eligible to answer this questionnaire.

Please be informed that this study is purely for academic purposes and that all information obtained shall be kept confidential.

Thank you.

**FORM NUMBER**……

**PART I**

## SECTION A: SOCIO-DEMOGRAPHIC CHARACTERISTICS OF HEALTHCARE PROFESSIONALS

Please tick {√) the response applicable to you.

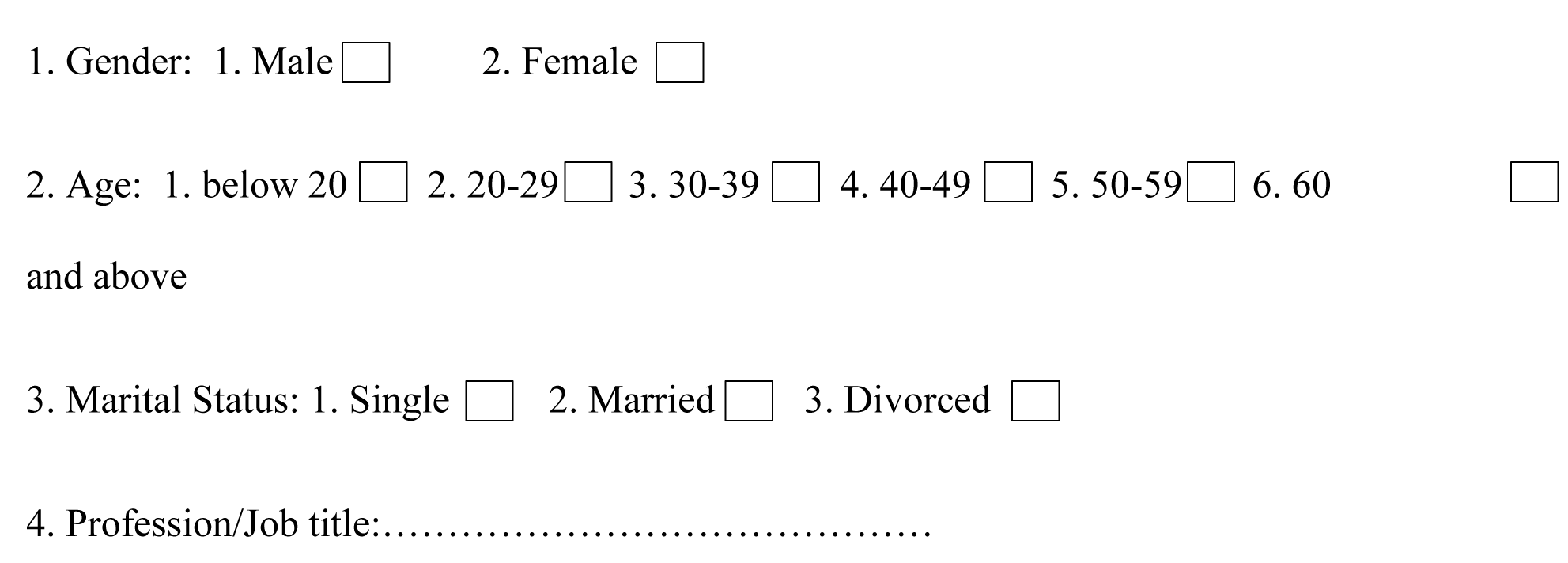
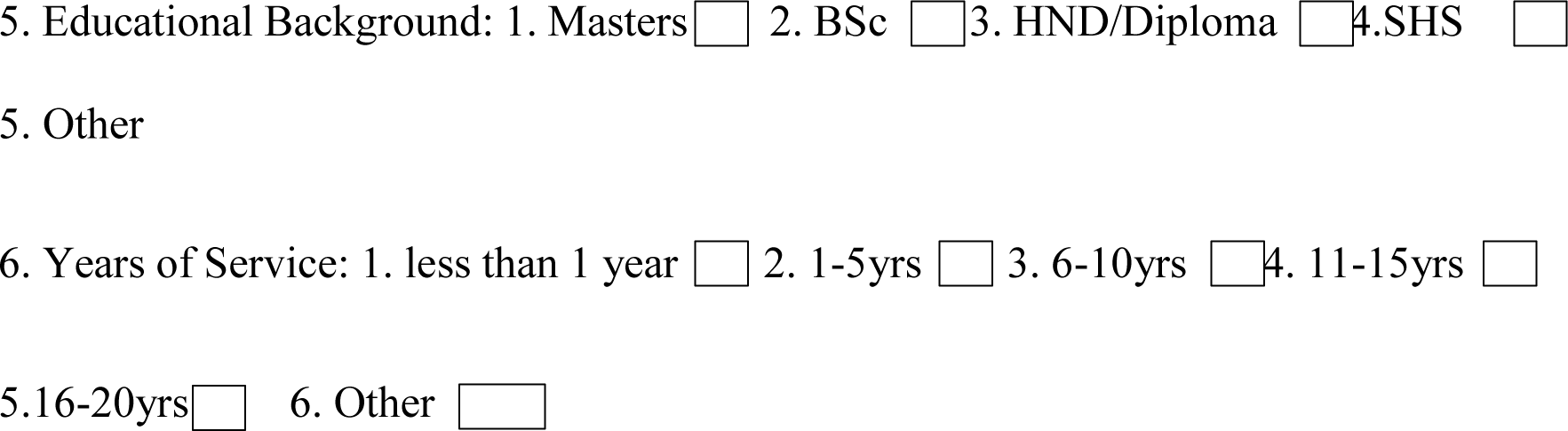

**PART II**

In section B below, talks about how to determine the impact of Health Information Technology on patient safety. The response to each statement is a True or False and rated on a scale of 1, 2, 3, 3, 4, and 5 respectively. Five (5) is the highest value on the scale; while 1 represents the lowest. Please tick (√) the response as applicable to you

## **B.** DETERMINING THE IMPACT OF HEALTH INFORMATION TECHNOLOGY ON PATIENT

| STATEMENT | RESPONSE ( please, tick(✓)) |  |  |  |  |
| --- | --- | --- | --- | --- | --- |
|  | 1 | 2 | 3 | 4 | 5 |
|  | Strongly Disagree | Disagree | Uncertain | Agree | Strongly Agree |
| 7. Health Information Technology is important in health care delivery |  |  |  |  |  |
| 8. Do you believe Health Information Technology has made operations in the hospital setting easier and faster? |  |  |  |  |  |
| 9. Has Health information Technology reduce medical errors? |  |  |  |  |  |
| 10. Health Information Technology has help to reduce patient waiting time |  |  |  |  |  |
| 11. Health Information Technology has help to transfer medical records of patients from one facility to the other in case of emergency. |  |  |  |  |  |

**PART III**

**In sections C to D below, the response to each statement is rated on a scale of 1, 2, 3, 4, and 5. Five (5) is the highest value on the scale; while 1 represents the lowest. Please tick (√) the response as applicable to you and the hospital**

## SECTION C: EXAMING THE USAGE OF HEALTH INFORMATION TECHNOLOGY ON PATIENT SAFETY

|  | RESPONSE ( please, tick(√)) |  |  |  |  |
| --- | --- | --- | --- | --- | --- |
| STATEMENT | 1 | 2 | 3 | 4 | 5 |
|  | Strongly Disagree | Disagree | Uncertain | Agree | Strongly Agree |
| 12. Health Information Technology has made it possible to identify and search for patient records without difficulty. |  |  |  |  |  |
| 13. Health Information Technology has helped patients to book appointment with their physician at home without necessarily walking to the facility. |  |  |  |  |  |
| 14. Health Information Technology such as Clinical Decision Support Systems (CDSS) has made it easy for Practitioners to make clinical decision when delivering care to patients. |  |  |  |  |  |
| 15. Patients are able to view their medical history from home by the use of the patient portal emerged from Health Information Technology. |  |  |  |  |  |

**PART IV**

## 1. SECTION D: THE DIFFICULTY IN USING HEALTH INFORMATION TECHNOLOGY TO PROVIDE CARE TO PATIENTS

**PART IV** **1. SECTION D: THE DIFFICULTY IN USING HEALTH INFORMATION TECHNOLOGY TO PROVIDE CARE TO PATIENTS**
| STATEMENT | RESPONSE ( please, tick(√)) |  |  |  |  |
| --- | --- | --- | --- | --- | --- |
|  | 1 | 2 | 3 | 4 | 5 |
|  | Strongly Disagree | Disagree | Uncertain | Agree | Strongly Agree |
| 16. Health Information Technology has made it possible to know the sex of an unborn child before it is born. |  |  |  |  |  |
| 17. Despite the importance of Health Information Technology, it also has drawbacks. |  |  |  |  |  |
| 18. Lack of training on the use of Health Information Technology is an issue to the users of it. |  |  |  |  |  |
| 19. When there's light out at places where there's no plant, a patient can lose his or her life. |  |  |  |  |  |
| 20. Health Information Technology which absolute or old in nature can bring a lot of errors. |  |  |  |  |  |
| 21. Personnel who are more skilled to use HIT in their department should be allowed to use it. |  |  |  |  |  |

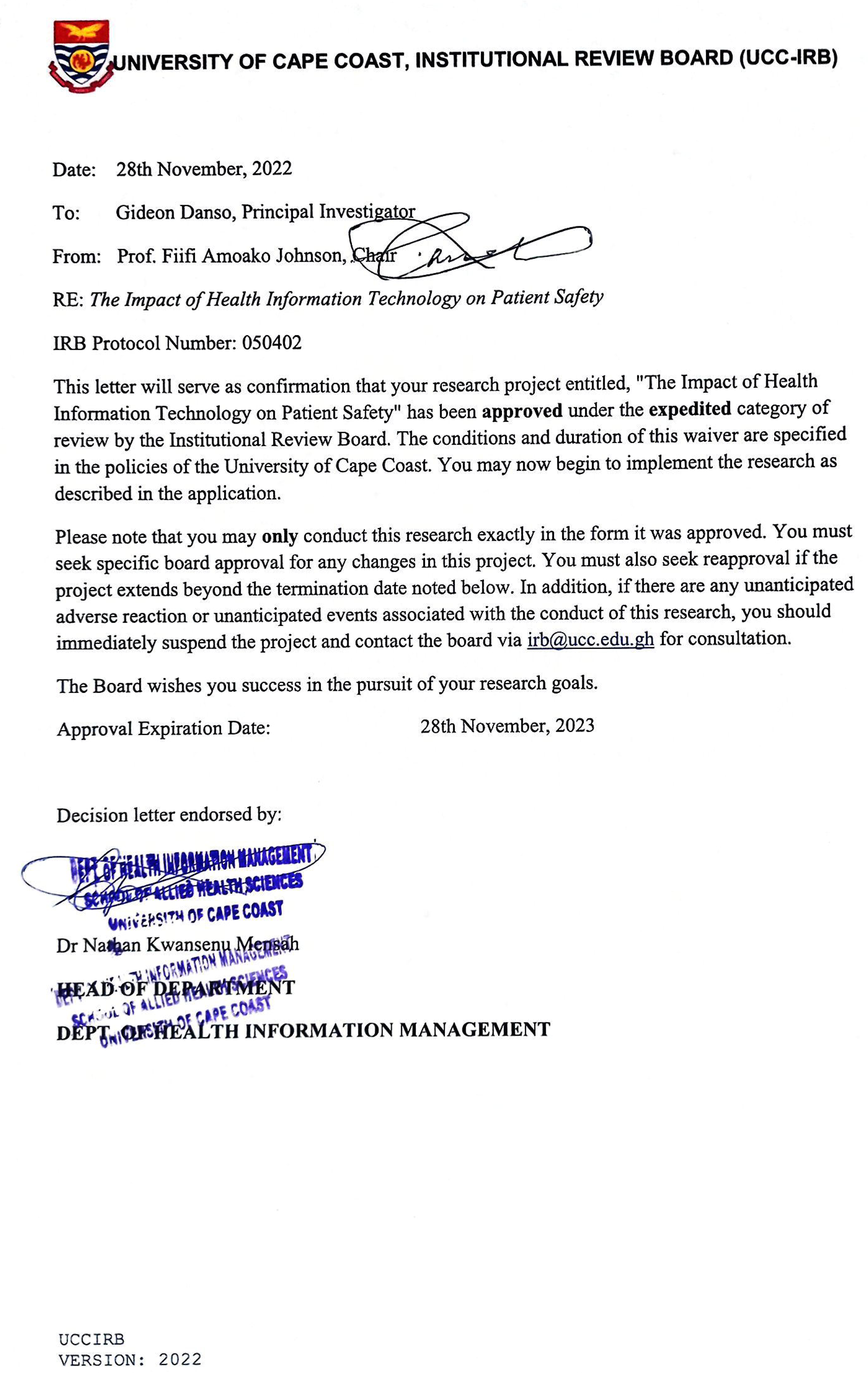

## Data Availability

All data produced in the present work are contained in the manuscript.

